# Graph-Based Patient Representation for Multimodal Clinical Data: Addressing Data Heterogeneity

**DOI:** 10.1101/2023.12.07.23299673

**Authors:** Suparna Ghanvatkar, Vaibhav Rajan

## Abstract

Patient data comprises of different modalities like clinical notes, lab results, and radiological investigations. Predictive modeling on this patient data is challenging due to the heterogeneity among patients and modalities captured, such as no ECG recordings for a few patients. Ensuring model applicability to all admitted patients requires addressing three key factors: i) handling modalities at disparate time-scales (e.g., once in a few hours medication and 125 Hz ECG need to be handled by same model), ii) handling missing modality (e.g., ECG not recorded), and iii) modeling temporal interactions between modalities over time (e.g., the ECG fluctuation triggered some lab test order). Existing literature often doesn’t simultaneously address these requirements. Therefore, we propose a novel patient representation approach inspired by clinical workflows, representing each patient as a graph. We categorize patient data into two main components: observations and care team actions, allowing cross-modality temporal interaction when actions consider the previous observations. We define observation nodes to capture modality-specific data within the time between two actions, and action nodes capture data of the actions undertaken, with edges to capture the temporal dependencies. To address missing modalities and time-scale disparities, we define node types for different modalities and use modality-specific representation for the nodes; implying a missing modality is equivalent to a missing node type in the graph. Aligned with clinical workflows, this patient-graph representation aims to enhance the practicality of predictive systems for various healthcare tasks, from mortality risk assessment to medication recommendations, thereby improving clinical decision support.

## 1 Introduction

Clinical Decision Support Systems (CDSS) have a fundamental goal: to enhance clinical decision-making by providing timely information. However, they often fall short of this goal, burdening clinicians with irrelevant or inaccurate predictions and disrupting their workflows [Sutton et al., 2020]. Despite the potential benefits of CDSS, these systems face significant challenges that demand attention. For example, an assessment of a proprietary sepsis prediction model in hospitals revealed significant delays in alerts, triggering them 68 to 145 minutes after clinicians’ actions. This highlights the existing systems’ limited ability to support timely decision-making. D’Hondt et al. [2022] delved into the barriers faced by ICU clinicians when using predictive CDSS, revealing predominantly technical challenges. These include varying patient measurements, insufficient data for different type of patients, and data silos between physical systems like bedside monitors and hospital Electronic Health Records (EHR) systems. Addressing these practical yet technical obstacles is essential to enhance the predictive performance of CDSS.

These challenges are inherently tied to the complexity of clinical data and its inherent heterogeneity. Given the practical challenges elucidated by D’Hondt et al. [2022], we identify two critical heterogeneities in clinical data that need to be addressed:

- Temporal Heterogeneity: Measurements at different time-scales but may influence one another over time (e.g., high-frequency ECG data at 125 Hz vs. laboratory tests at once in couple of hours frequency, where the lab tests may be influenced by the ECG data)
- Measurement Heterogeneity: Varied measurements across patients leading to missing modalities (e.g., a patient may have no X-ray data as it may not be needed for diagnosis).

The measurement heterogeneity implies that predictive systems must model patients with any modality missing. For example, one patient may undergo ECG and lab tests, while another, admitted with a different condition, may have X-rays and microbial cultures captured. Predictive systems relying on specific modalities cannot accurately predict both patients.

A simple approach to handling disparate time-scales and missing modalities is to use a late-fusion model to integrate the modality-specific representations of the different modalities before performing prediction (e.g., Soenksen et al. [2022]). However, this approach lacks the capability for cross-modality interactions over time, which are crucial in aligning models with clinical workflows. Clinicians base decisions on an integrated view of patients, such as ordering specific laboratory investigations based on observed changes in the ECG. Therefore, to ensure applicability across all patients and alignment with clinical practice, the following challenges need to be handled: i) managing disparate time-scale modalities (e.g., periodic medication and high-frequency ECG), ii) modeling interactions between these modalities over time (e.g., ECG fluctuations triggering lab test orders), iii) handling missing modalities (e.g., absent ECG recordings), and iv) no requirement of specific modality (i.e. can handle two patients with disjoint modalities captured like one patient with ECG and vitals and another with X-Ray and laboratory procedures). The first two are a result of the temporal nature of the clinical data, and the latter two are implications of measurement heterogeneity.

The extant literature on multimodal data often cannot model these four challenges simultaneously. Current patient risk prediction models often handle multimodal data at similar time resolutions, such as lab tests or clinical notes [Kline et al., 2022]. While high-frequency time series like ECG have been studied independently for risk prediction, very few works integrate them with sparse EHR time series like lab tests (e.g., Xu et al. [2018]). However, these methods often cannot handle missing modalities. Although some research addresses data imputation within modalities, these methods do not apply to missing modalities [Zhang et al., 2022]. As emphasized in our example and D’Hondt et al. [2022], these challenges mirror real-world scenarios and are significant barriers to clinicians’ adoption of CDSS. So, addressing these challenges simultaneously is crucial to handle the two clinical data heterogeneities.

Thus, we define our research objective as: *To develop a modeling strategy for multimodal predictive systems that accounts for the challenges due to the temporal and measurement heterogeneity in the clinical data*.

In this paper, we develop a patient representation strategy that satisfies our research objective. Our approach considers patient data as sequences of clinician-triggered actions that delineate observed data, thereby providing a faithful representation of their medical journey. To capture temporal dependencies and other related connections between the actions and observed data, such as diagnoses based on confirmatory lab tests, we adopt a graph-based model. In this representation, the actions and the observations are depicted as nodes, and edges connect these nodes to capture temporal dependency and other connections. The temporal dependency captured through edges between the observations that preceded and influenced the action allows for cross-modal temporal interactions. Additionally, we employ modality-specific models for observations within each distinct data modality, accommodating diverse time scales present in clinical data. Representation as a graph also allows us to handle missing modalities without requiring the presence of any particular modality, allowing for disjoint modalities between patients; thus handling measurement heterogeneity. We evaluate our approach on sepsis prediction among Intensive Care Unit (ICU) patients and get encouraging initial results. Consequently, the next sections will first discuss gaps in the existing literature, followed by a detailed presentation of our methodology, experimental setup, and initial findings. Finally, we will discuss the future implications and plans for our approach.

## 2 Related Work

The high-frequency time-series, notably electrocardiogram (ECG), have been independently studied and used in several prediction models [Angelotti et al., 2018]. Angelotti et al. [2018] use ECG and arterial blood pressure (ABP) signals to train classifiers like SVM, KNN, and decision trees to predict acute hypotensive episodes in ICUs. The information in these signals is also found to strongly correlate to the mortality of patients [Raghunath et al., 2020]. However, the integration of these high-frequency time-series data with sparse clinical time-series extracted from Electronic Health Records (EHR), such as intermittently captured laboratory values, remains limited, as noted by a recent review of deep learning methods for using physiological signals in clinical predictions [Morid et al., 2023]. Xu et al. [2018] develops an architecture for addressing this temporal heterogeneity by applying it to decompensation prediction. However, it requires the presence of the ECG signal, thus not modeling the measurement heterogeneity. Thus, there is limited literature that allows modeling disparate timescales of data, while allowing for missing modalities.

Often, the models utilizing multimodal clinical data just concatenate different modality representations [Huang et al., 2020]. Certain models, instead of combining modalities, commonly designate a ‘primary’ modality required for predicting outcomes, mandating its presence for all patients within the model’s scope. Consequently, the models cannot predict for patients lacking this primary modality. For instance, in RAIM [Xu et al., 2018], the ECG signal is required, and the presence or absence of laboratory results is handled. Similarly, most of the models have such a requirement, implying they do not handle the measurement heterogeneity (e.g. Rajkomar et al. [2018]). However, as noted earlier, in real-life scenarios, different patients have different modalities recorded, and the specific modality recorded could be sufficient to capture the patient’s status. Thus, requiring a primary modality is restrictive.

To address missing modalities in patient data we can draw insights from literature on missing values. These very effectively handle the within-modality missing values, such as using Gaussian Process for missing value imputation [Futoma et al., 2017]. However, these strategies often do not extend when the entire modality is missing, i.e., when there is no observation for the particular modality. Some studies opt to exclude patients lacking specific modalities (e.g., Xu et al. [2018], Rajkomar et al. [2018]). Recent advancements use generative models to tackle missing modalities (e.g., Pan et al. [2022]). Yet, these models may introduce noise by assuming an ambiguous latent space for clinical data [Zhang et al., 2022]. Zhang et al. [2022] propose identifying similar patients based on available modalities to impute missing modalities in the latent space directly, addressing cases where modalities are missing due to system errors or noise. However, it overlooks systematic biases in patient data, contributing to measurement heterogeneity, such as the absence of X-ray for patients who may not require it for their diagnosis. Thus, while these studies address measurement heterogeneity, they primarily focus on handling modality absence due to errors rather than systematic decision-based biases in the data.

In recent years, Graph Neural Networks (GNNs) have gained significant attention and demonstrated promising results in various domains, including clinical prediction tasks. GNNs are a class of deep learning models designed to operate on graph-structured data. Clinical prediction tasks involve utilizing patient data to make accurate predictions about disease diagnosis, prognosis, treatment response, and other healthcare outcomes. One stream of studies uses entire data or set of patients represented as graphs and these representions are used for downstream tasks. For example, Zhang et al. [2022] find the similar patients by representing patients by their profiles and use for further downstream task of missing modality imputation. A common approach of application of GNN to clinical context is creating a large heterogeneous network with relations between patients and their associated information, such as medications or diagnosis. For a new patient, the prediction tasks are reframed as node classification or link prediction tasks, depending on the structure of the graph. For example, Tang et al. [2023] create network of the admissions of patients based on logitudinal EHR information. For predicting the readmission, node clasification is done to predict if the patient (represented as node) will be readmitted. Readers can refer to Lu and Uddin [2023] for comprehensive review on GNN approaches for disease prediction. Additionally, another stream of applications use GNNs along with knowledge graph to perform knowledge-driven predictions. For example, Ye et al. [2021] uses information from knowledge graphs to obtain possible disease progression and combines with predictions from EHR to perform knowledge-based risk-prediction. Thus, though the GNN approaches have been developed for generating predictions, none of them use graphs to address all the four challenges we have identified.

In summary, despite advances in multimodal prediction, clinical applications often overlook clinical data heterogeneity. Notably, current studies lack comprehensive solutions addressing both temporal and measurement heterogeneity while enabling inter-modality interactions over time. These crucial factors - i) managing disparate time-scale modalities, ii) modeling temporal interactions between these modalities over time, iii) handling missing modalities, and iv) no requirement of specific modality - are essential in patient prediction. However, the current literature falls short in simultaneously addressing these requirements.

## 3 Method

### 3.1 Overview

To tackle challenges arising from clinical data heterogeneity, we design a graph-based patient representation method. Nodes within these graphs correspond to patient EHR data from distinct modalities within specified timeframes, while edges denote temporal actions and causal links, including symptom-to-intervention connections. For example, an edge can be used to represent if patient symptoms led to a diagnosis or intervention. However, adopting a fixed time-frame, such as one hour per node, could lead to larger graphs for patients with extended hospital stays, potentially causing computational complexities. Determining an optimal time-frame (e.g., one hour versus half an hour) involves a trade-off between computational efficiency and capturing meaningful temporal relationships. To address this, we opt for a ‘dynamic’ approach in determining these time-frames instead of relying on predefined static intervals, as elaborated further in this section.

Our approach to dynamic time-frames draws inspiration from clinical workflows. When a patient is admitted to the hospital, clinicians initiate a sequence of observations followed by subsequent actions, which are further followed by additional observations and actions, forming a chain of events until discharge. So, the patient data can be viewed as a sequence of actions (e.g., medical procedures, medications) and observations (e.g., vitals, pain scores) done by the care team. We designate the timings of clinicians’ actions as dynamic time-points. This approach allows for temporal interaction between different data modalities—when clinicians take actions, they consider preceding actions and observations, enabling interactions across different modalities.

Let us illustrate our method using the example in Figure 1. The patient has different modalities of data, where the different outlines denote different modalities. The yellow and red nodes represent observation and action nodes, respectively. The observations consist of all the data captured, like the vitals observed, lab results obtained, the image output, etc. The action node contains the information of the actions performed by the care team, such as prescriptions, lab test orders, etc. We have three types of edges for this example patient: i) temporal dependence, ii) observations associated to an action, and iii) delayed observations of actions. The last two edge types connect actions to observations; however, the latter captures a delayed result, which may be obtained by lab test order. The second edge type captures the observations recorded along with the action, such as the medications given with the prescription or observations from physical examination of patient.

**Figure 1:**
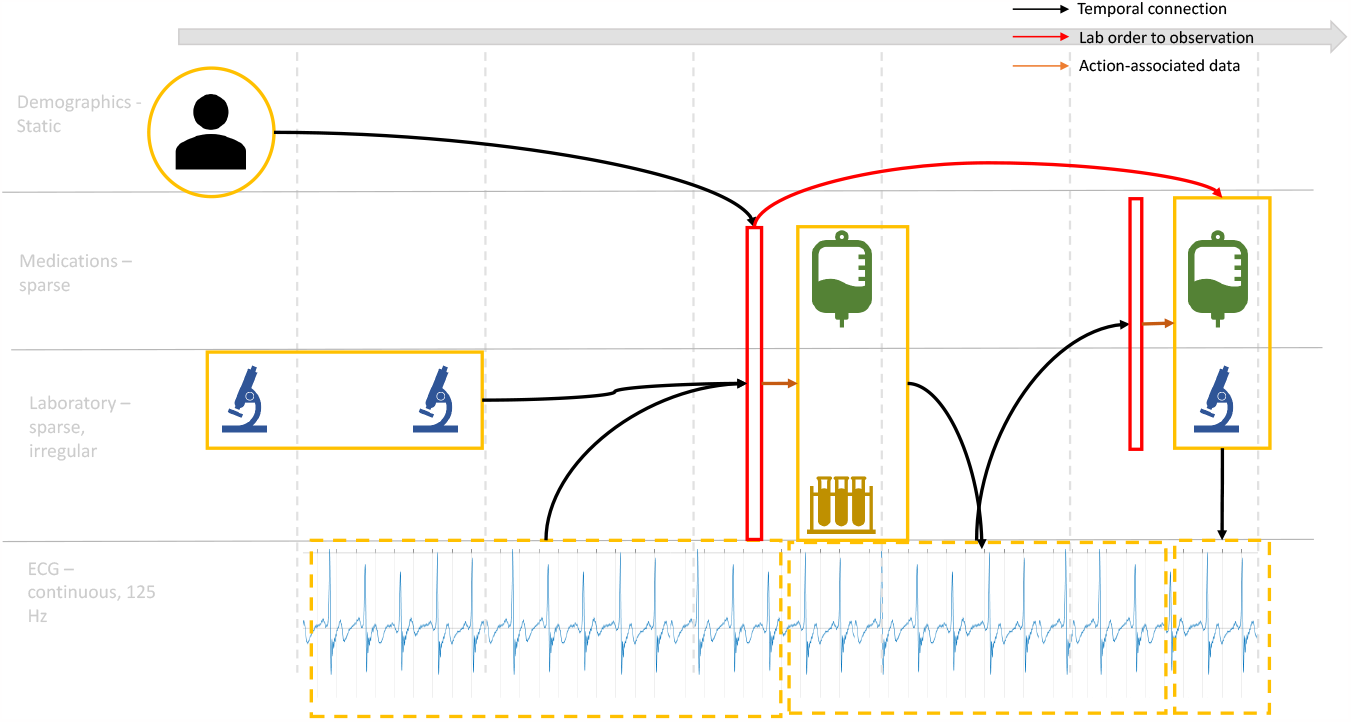
Example representation of patient as a graph. The red indicate actions like prescription, and yellow indicates observations like laboratory investigation result. The various outlines indicate different independent models used to obtain the node representations. These nodes may be connected through edges indicating their temporal dependence (black edges) as well as specific edges, such as those based on causation obtained from clinical notes or from the fact that laboratory records were ordered and action was followed after their reports.

Our approach handles all the four challenges outlined earlier. Firstly, due to modality-specific nodes, the representations can be obtained using modality-specific models, allowing disparate time scales of data. Second, inspired by clinical workflows, we allow cross-modality interaction during the timings of the actions. Third, the graph-based approach inherently handles measurement heterogeneity: the graph structure allows for a variable number of nodes. Therefore, any modality can be missing for any patient, and this missingness is indirectly captured in the graph representation by the absence of certain modality nodes. Lastly, the graph-based approach ensures that we do not have a requirement of any specific modality as long as that modality has been observed in the training data as well.

We now describe our method in detail by splitting it into three steps: Data segmentation, Graph representation, and Graph classification.

### 3.2 Data segmentation

As mentioned earlier, patient data can be viewed as a sequence of observation and actions, where the observations are captured until an action is taken. Thus, to generate patient graph representation, the first step is to determine the observation and action segments. To determine these *segments* of data, we categorize the records in patient data as either actions or observations. Action data represents actions performed by doctors, such as ordering laboratory measurements or conducting medical procedures. Observations capture recorded data, including routine vitals, pain measurements, medications given, etc. The action segment comprises a series of actions occurring within a defined time interval of each other, denoted by *δ*. The observation segment is delineated by action timings, implying all observations before the start of the next action segment and after the end of the previous action segment are part of the observation segment. These segmented observations and action data form the basis for the nodes within the patient graph.

These data segments, representing the nodes within our patient graph, are interconnected by edges to establish temporal dependencies. Clinicians often consider previous observations when determining subsequent actions, forming the basis for these temporal connections. Additionally, connections between segments can include actions linked to other observations or action data. For instance, connections between segments are formed when a specific action, like prescribing medications, is in anticipation of a subsequent action, such as a medical procedure. These inter-segment connections illustrate the sequential relationship between actions and observations within the patient’s care journey, contributing to the structure of our patient graph representation.

### 3.3 Graph Representations

In our methodology, we impose a restriction that each node can be associated with only one modality of data. This implies that if, in an observation segment, we have an ECG recording along with text notes, then we will have two observation nodes resulting from the segment: one for ECG (high-frequency time-series) and the other for text. Doing so will allow us to handle both, temporal and measurement heterogeneity in the data. Thus, we will have different node types based on the modalities in the dataset and whether that node is an observation or action node. The different edges determined in the previous steps will connect every resulting node from the segments of actions and observations. Thus, we will get a heterogeneous graph having different node types and edge types.

Let us define a heterogeneous graph that is obtained to represent the patient as *G* = (*V, E*), where *V* is the set of nodes of dimension *d* with each node associated with a node type and *E* is the set of directed edges with each edge associated to an edge type. To obtain the nodes for graph form, we use modality-specific models for the node representations. Thus, ECG data is converted to a *d* dimensional vector representation using a model trained on ECG data for generating embeddings.

### 3.4 Graph Classification

The heterogeneous graph representing a patient that is generated in the previous step is used to train a Graph Neural Network (GNN) for the prediction label for each patient. Thus, the graph classification step using GNN takes the *G* input and produces label *y* for the prediction task. The distinctive aspect of GNNs, such as Graph Convolution Networks (GCN), lies in their training methodology. Unlike traditional neural networks, where layers primarily focus on learning high-level abstractions, each layer of a GNN updates the node representations. As our input is a heterogeneous graph, this strategy needs to be adapted for the training task. A common approach is to consider a homogeneous graph consisting of only one edge type and aggregate the updates obtained for each node from these edge-specific homogeneous graphs. Thus, the different node and edge types in our graph *G* need to be converted to unique ‘connection’ consisting of edge and node type combinations existing in the graph. Thus, we get graph *G* = (*V, R*), where *V* is a set of node vectors with dimension *d* and *R* is a set of relations or connections between these nodes, with the relation type based on the unique combination of node and edge of its original edge.

At each layer of the HeteroGCN, we begin by using the node representations, denoted by *h*^(*l*)^. Let *N* represent the total number of nodes, and *D*^(*l*)^ represent the dimensionality of the node features at layer *l*.

This information undergoes a transformation to update the representation of each node for the next layer. Thus, the updated representation of node *v* at the next layer is calculated using formula 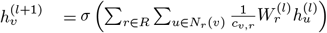 Here, 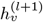 is the representation of node v at layer l+1, *σ* is an activation function (e.g., ReLU), R is the set of edge types, *N*_*r*(*v*)_ represents the neighbors of node v connected by edges of type r, *c*_*v,r*_ is a normalization constant, and 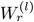 is a learnable weight matrix associated with edge type r at layer l.

In simpler terms, within HeteroGCN, individual graphs are constructed for each distinct relation type. For these relation-specific graphs, nodes are updated using the GCN method, tailored for homogeneous nodes and edges. The resulting node representations from these relation-specific graphs are then aggregated and processed through a non-linear transformation, such as the *σ* activation function (like ReLU), to generate updated representation.

The HeteroGCN model updates the node representations by stacking multiple graph convolutional layers. Each layer refines the node representations by incorporating information from neighbors and their respective edge types. These updated node representations are combined using various readout strategies, such as the mean of all updated node vectors or attention mechanisms over all updated node vectors, to generate a patient-level representation. Subsequently, a neural network is trained end-to-end to predict the label *y* based on the obtained patient representation.

In summary, unlike the extant literature, our approach addresses the four challenges simultaneously, while building on existing work. Our approach successfully handles the temporal and measurement heterogeneities present in medical data by utilizing modality-specific models and the flexibility of the graph structure.

## 4 Experiment

### 4.1 Data

We use the publicly available EHR dataset, MIMIC-IV [Johnson et al., 2023b], containing deintendified clinical data from Beth Israel Deaconess Medical Center (BIDMC). It contains information for all patients admitted to BIDMC emergency or ICU between 2008-2019. The data contains a total of roughly 70,000 ICU stays. Discharge summaries and radiology report notes are available for these patients as a part of the MIMIC-IV-Note project [Johnson et al., 2023a]. For a subset of patients, Chest X-ray images associated with all their hospital stays are available as part of the MIMIC-IV-CXR project [Johnson et al., 2019]. The ECG data for a subset of these patients will also be soon released as a part of the MIMIC-IV Waveform project [Moody et al., 2022].

To create train, validation, and test splits for the dataset, we follow a stratified approach. Though we need to model the measurement heterogeneity, we also need to ensure that in train as well as test set, for every modality, at least one patient with that modality is present. We use three sources of data - chartevents, notes, and X-rays. Patients with duplicate admissions, i.e., the patients who were transferred, were merged into one record representing a patient’s hospital stay. Patients need to have at least one source of input. That leaves us with 18389 patient stays in the train set and 4571 in the validation set.

### 4.2 Task

We consider an early warning system for the detection of the onset of sepsis in ICUs. Sepsis is a life-threatening medical emergency that has a high mortality rate; prompt treatment can improve the chances of a successful outcome [Paoli et al., Singer et al.]. It is an organ dysfunction that is often managed in hospitals using the Sequential Organ Failure Assessment (SOFA) score, which is based on specific criteria within each system, such as blood pressure, platelet count, and level of consciousness [Singer et al.]. The system alerts when a patient is predicted to develop sepsis in the next 6 hours (prediction). So, we evaluate our method on the binary prediction task of whether a patient will develop sepsis or not in the next 6 hours.

The labels and timing for the sepsis are determined by using the earliest time at which the patient had SOFA >= 2, and clinicians suspect an infection. The time of suspicion of infection is calculated as the earliest between the culture time and antibiotic time, which are the approaches used to confirm and start treatment for steps. To obtain the time of suspicion of infection and sepsis label, we use mimic-code repository [Johnson et al., 2018]. The length of patient stay is different for each patient in our experiment setup. As we want a system that predicts the onset of sepsis before doctors suspect it, we set the target time as 6 hours before this time of sepsis. For the patients who do not have sepsis, the target time of prediction was 48 hours. Thus, the input is a variable-length patient stay until the target time when we want the prediction, and the output is a binary label indicating if the patient will develop sepsis in the next 6 hours or not.

### 4.3 Experiment Setup

#### 4.3.1 Data segmentation

We use the label column of the chart events (e.g., non-invasive blood pressure) for a patient to determine if an event must be considered action or not. All laboratory investigation orders and procedures are considered actions. All routine vitals measured for patients are considered observations. For the other categories of events, such as neurological or cardiovascular, we perform the classification based on examples obtained from GPT-3.5 LLM. The prompt used to obtain the classification is:

~~~
I want to classify a list of actions from patient chart from EHR into observations and
actions by doctors. For example, routine vital signs or pain level assessment, etc. would
be observations. Action would be medications, access lines, alarm settings, procedures
or lab measurement ordering. The list I want to classify is this:
~~~

We use two types of edges: i) the temporal connection and ii) the delayed result of lab tests. For our experiment, we add the observation data captured along with the actions, i.e., the various notes, symptoms, medication records, etc, as part of the action segment. All actions (along with associated observed data) within 10 minutes of one another are made part of the same action segment. The timing of these actions provides us with the timings used to determine the observation segments.

#### 4.3.2 Graph Representations

We use three modalities: Numeric time series, Text, and Images. The chartevents that have text data associated with it, such as symptom observation, are classified as text modality. Nodes in our graph representation are modality-specific; hence, nodes associated with actions accompanied by text data are categorized differently from those accompanied by numeric data. Similarly, we distinguish three types of observation nodes.

For the Numeric time-series modality, our representation involves creating matrices wherein the rows represent various measurements (e.g., Creatinine levels), and the columns denote the maximum number of hours within an observation segment. Each matrix entry captures the recorded data value of the measurement (row) at a particular hour since the beginning of the observation segment (column). To learn node representations, we use an autoencoder trained on these matrices, generating node representations of 100 dimensions.

For the text and image modality, we follow the same preprocessing and embedding strategy as Soenksen et al. [2022]. For Text modality, we consolidate all text data within a segment into a single text note. Subsequently, we employ the Clinical BERT pre-trained model [Alsentzer et al., 2019] to obtain embeddings for these notes. For the X-ray data, we use TorchXRayVision package [Cohen et al., 2022] with a model trained on CheXpert dataset [Irvin et al., 2019]. This model provides embeddings based on prediction probabilities for 18 diseases the model was trained on, such as Pneumothorax or Edema.

#### 4.3.3 Graph Classification

The unique combination of the (source node type, edge type, destination node type) is used to determine the ‘connections’ for the heterogenous graph representation of the patient. For the task of graph classification, we use HeteroGCN using the DGL library. The patient vector representation is obtained through an average readout, which takes an average of all the updated node vector representations.

### 4.4 Experimental Results

On performing data stratification, we get train and validation split of 18389 and 4571 patients admissions, respectively. The label used is onset of sepsis in next 6 hours. The patients that do not have a single ‘action’ node, based on the classification of action, have been dropped resulting in 18344 train and 4560 patients. Out of these patients, only 2918 patients of train set and 713 patients of validation set have x-rays. Similarly, only 14427 train patients and 3609 validation patients have radiology notes. The results are presented for classification of the 4560 samples in Table 1.

**Table 1:**
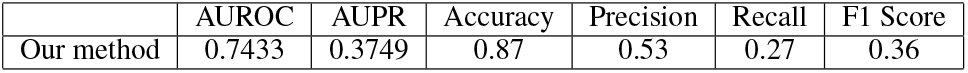
Results on our Experiment Setup.

|  | AUROC | AUPR | Accuracy | Precision | Recall | F1 Score |
| --- | --- | --- | --- | --- | --- | --- |
| Our method | 0.7433 | 0.3749 | 0.87 | 0.53 | 0.27 | 0.36 |

To compare the predictive performance of our system, we plan to compare with three other baselines: i) HAIM [Soenksen et al., 2022]: uses late-fusion of modality-specific representations, ii) M3Care [Zhang et al., 2022]: missing modality imputation in latent space using similar patients, and iii) ARMOUR [Liu et al., 2023]: missing modality imputation using transformer based model.

## 5 Discussion

We introduce a novel patient-representation approach that improves the practicality of predictive systems by allowing prediction for all patients, regardless of the modality captured for the patient. Our representation strategy allows us to handle both the critical heterogeneities observed in the data, namely temporal and measurement heterogeneity. Our approach can accommodate patients with different durations of stay, as it allows for the flexible incorporation of different types of measurements and their varying availability. By leveraging the inherent flexibility of the graph structure, we can effectively handle the heterogeneity in measurements across different patients. Using the modality-specific models allows us to model temporal heterogeneity while ensuring the system can improve with updated research for each modality. Our approach aligns with clinicians’ workflow, making it potentially accessible for explaining predictions to medical professionals.

However, our experiment design involves several considerations, including action and observation classification and modality-specific models. Future experiments will explore alternative design choices, particularly refining action classification to accurately capture care team interventions and incorporating currently missed patient actions. Additionally, different node representation strategies, especially for the numeric modality, and variations in GNN modeling and readout strategies will be explored to optimize hyperparameters for patient prediction.

Moreover, our graph-based representation holds promise for integration with knowledge graphs, enabling personalized precision health applications by incorporating patient-specific relations as edges, such as allergies and medical history. Beyond healthcare, our method’s applicability extends to various domains dealing with multiple data sources, facilitating interactions between modalities over time while addressing missing modality issues.

## Data Availability

All data used are available online with credentialed access

https://mimic.mit.edu/

